# The contributions of cartilage endplate composition and vertebral bone marrow fat to intervertebral disc degeneration in patients with chronic low back pain

**DOI:** 10.1101/2021.10.25.21265485

**Authors:** Noah B. Bonnheim, Linshanshan Wang, Ann A. Lazar, Jiamin Zhou, Ravi Chachad, Nico Sollmann, Xiaojie Guo, Claudia Iriondo, Conor O’Neill, Jeffery C. Lotz, Thomas M. Link, Roland Krug, Aaron J. Fields

## Abstract

**Purpose:** The composition of the subchondral bone marrow and cartilage endplate (CEP) could affect intervertebral disc health by influencing vertebral perfusion and nutrient diffusion. However, the relative contributions of these factors to disc degeneration in patients with chronic low back pain (cLBP) have not been quantified. The goal of this study was to use compositional biomarkers derived from quantitative MRI to establish how CEP composition (surrogate for permeability) and vertebral bone marrow fat fraction (BMFF, surrogate for perfusion) relate to disc degeneration.

**Methods:** MRI data from 60 patients with cLBP were included in this prospective observational study (28 female, 32 male; age = 40.0 ± 11.9 years, 19–65 [mean ± SD, min–max]). Ultra-short echo-time MRI was used to calculate CEP T2* relaxation times (reflecting biochemical composition), water-fat MRI was used to calculate vertebral BMFF, and T1ρ MRI was used to calculate T1ρ relaxation times in the nucleus pulposus (NP T1ρ, reflecting proteoglycan content and degenerative grade). Univariate linear regression was used to assess the independent effects of CEP T2* and vertebral BMFF on NP T1ρ. Mixed effects multivariable linear regression accounting for age, sex, and BMI was used to assess the combined relationship between variables.

**Results:** CEP T2* and vertebral BMFF were independently associated with NP T1ρ (*p* = 0.003 and 0.0001, respectively). After adjusting for age, sex, and BMI, NP T1ρ remained significantly associated with CEP T2* (*p* = 0.0001) but not vertebral BMFF (*p =* 0.43).

**Conclusion:** Poor CEP composition may play a significant role in disc degeneration severity and can affect disc health both with and without deficits in vertebral perfusion.

## Introduction

While intervertebral disc degeneration is prevalent in people without chronic low back pain (cLBP), there is a strong implication that disc degeneration causes cLBP in certain subgroups [1] and consequently, therapeutically slowing or reversing disc degeneration is a common but as yet unrealized clinical goal. Cell-based biologic therapies to regenerate the disc have demonstrated mixed efficacy in clinical trials, and a key obstacle to successful translation of these therapies is an incomplete understanding of the mechanisms of cellular homeostasis and the factors disrupting homeostasis in degenerating discs [2,3]. One specific knowledge gap is the uncertain role of cartilage endplate (CEP) composition relative to vertebral vascularity. Both endogenous and implanted cells inside the nucleus pulposus (NP) are nourished by glucose and oxygen diffusing from the vertebral capillary bed, across the CEP, and into the NP [4]. *In vitro* experiments demonstrate that physiologic fluctuations in CEP composition (affecting its permeability), as well as endplate-adjacent vascularity (providing the reservoir of available nutrients), can impact NP cell viability and function [4,5]. However, despite these insights, the contributions of CEP composition and vertebral vascularity to lumbar disc degeneration in humans is unknown. Identifying the relative contributions of these two factors could clarify the etiology of disc degeneration and point to novel strategies for assessing and/or enhancing the disc’s regenerative potential.

Advances in magnetic resonance imaging (MRI) enable quantitative assessment of the biochemical compositions of the CEP, vertebral bone marrow, and disc. Specifically, ultra-short echo-time (UTE) imaging enables measurement of CEP T2* relaxation times, which correlate with glycosaminoglycan (GAG) content, water content, and collagen-to-GAG ratios in the CEP [6]; chemical shift encoding-based water-fat MRI (CSE-MRI) enables assessment of bone marrow fat fraction (BMFF), which is inversely proportional to the amount of hematopoietic marrow and provides a proxy for perfusion [7–10]; and T1ρ imaging enables measurement of NP T1ρ relaxation times, which correlate with proteoglycan content and provide objective, continuous measurements of disc degeneration severity [11]. These MRI techniques have the potential to reveal factors influencing pathologic changes to the disc-endplate complex *in vivo*, but have not been previously combined in a single study of disc degeneration. Our goal was to integrate these advanced quantitative MRI techniques into a comprehensive study of CEP and vertebral bone marrow composition in relation to disc degeneration in cLBP patients. We hypothesized that CEP and vertebral bone marrow composition associate with disc health after adjusting for age, sex, and body mass index (BMI).

## Methods

### Subjects

Between January 2020 and August 2021, 84 patients with cLBP were prospectively recruited and imaged for this IRB-approved study. Written informed consent was obtained from each patient. All patients met the criteria for cLBP as defined by the National Institutes of Health Pain Consortium Research Task Force [12], assessed using the following two questions: (1) “How long has low-back pain been an ongoing problem for you?” and (2) “How often has low-back pain been an ongoing problem for you over the past six months?”. A response of greater than three months to question one, and a response of “at least half the days in the past six months” to question two, met the cLBP criteria. Major exclusion criteria were a history of lumbar spinal surgery or fracture, known disc herniation, autoimmune disorders (including ankylosing spondylitis, rheumatoid arthritis, and psoriatic arthritis), or malignancy. All recruitment and imaging was conducted at a single institution. Patient-reported measures for disability and pain were collected using the Oswestry Disability Index (ODI) and Visual Analog Scale (VAS), respectively.

### MRI

MRI of the lumbar spine was performed on a GE 3T Discovery MR750 scanner using an 8-channel phased-array spine coil (GE Healthcare, Waukesha, WI). MRI data used for the purposes of this study were acquired using sagittal acquisitions from a multi-echo UTE Cones sequence [13], a CSE-MRI sequence [7], a T1ρ mapping sequence [11], and standard clinical fast spin-echo sequences with T1- and T2-weighting (see Supplemental Content).

### Image analysis

To assess CEP composition, all four CEPs at the L4/5 and L5/S1 levels were manually segmented on the first UTE echo by a single trained annotator using a custom segmentation tool (IDL 8.8). The hyperintense CEP signal from the UTE images guided segmentation (Figure 1, top row). The same annotator re-segmented a subset of 20 CEPs from five patients a minimum of four weeks after initial segmentation to assess intra-rater reliability (intraclass correlation coefficient [ICC] for resulting T2* values = 0.91). After segmentation, T2* relaxation times were calculated and the 3D voxel data was rotated into a standard coordinate system (MATLAB R2020b) before a kidney-bean-shaped template was applied using methods described previously [14]. This algorithmic approach enabled consistent identification of the central CEP region adjacent to the NP. To account for the sensitivity of CEP T2* values to CEP orientation within the MRI bore, only CEPs oriented within ± 15° of the ‘magic angle’ (54.7°) in the sagittal plane were included, since T2* values are artificially depressed at other angles [6]. Subjects with CEP damage precluding segmentation were also excluded.

**Fig. 1.**
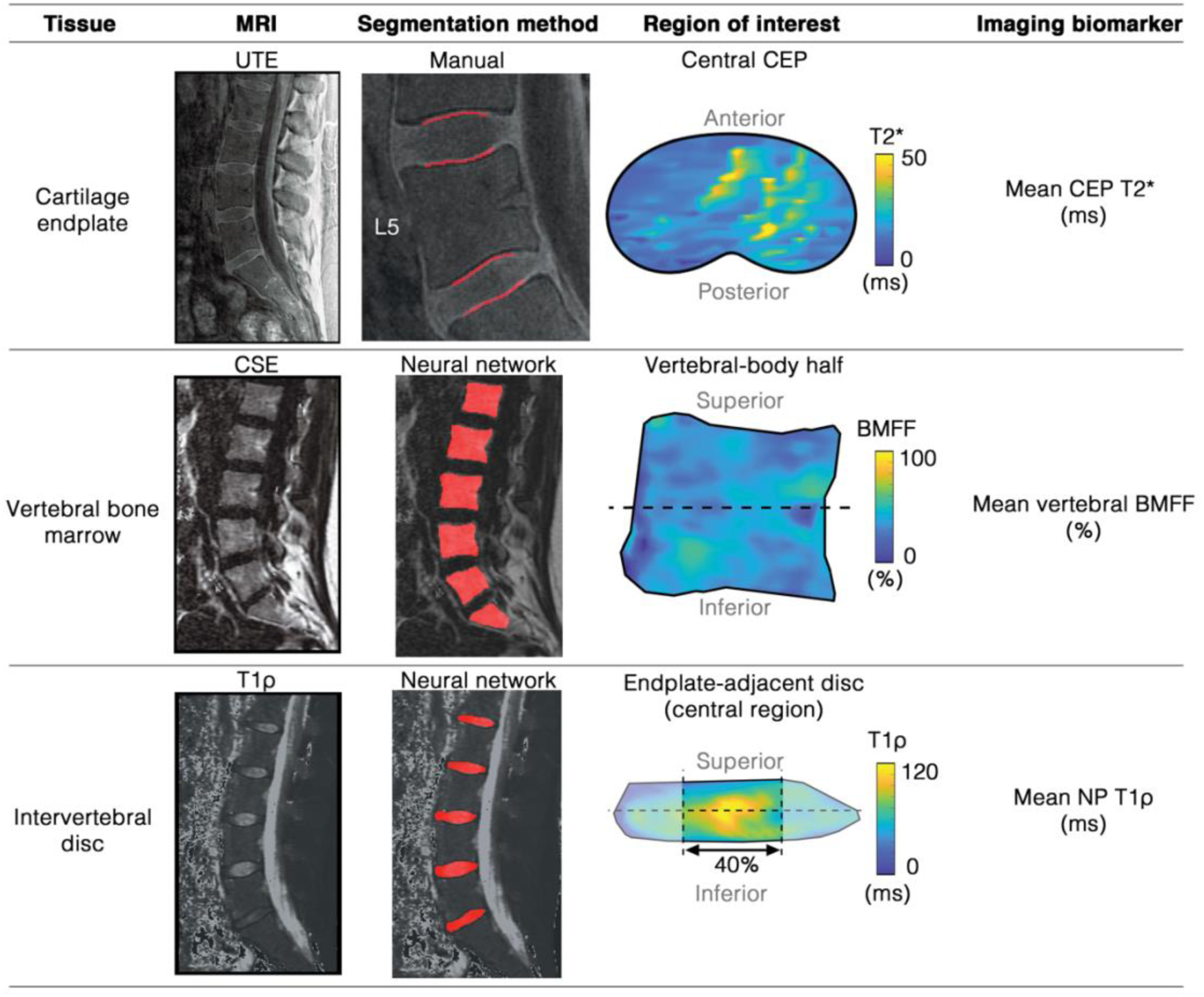
Representative MRI image, tissue segmentations, and biomarker visualization for the cartilage endplate (top row), vertebral bone marrow (middle row), and intervertebral disc (bottom row). CEP = cartilage endplate, BMFF = bone marrow fat fraction, NP = nucleus pulposus

To assess vertebral bone marrow composition, the L4, L5, and S1 vertebral bodies were segmented from five mid-sagittal slices of the CSE-MRI fat-fraction images using a neural network (Figure 1, middle row), applying manual corrections as needed [15]. The segmented 3D voxel data were rotated about the shape’s principal axes into a standard coordinate system. This approach enabled bisection of the vertebral body in order to isolate the hemi-vertebral region adjacent to each endplate.

To assess disc composition, the lumbar discs were automatically segmented from five mid-sagittal T1ρ images using a neural network (Figure 1, lower row) [16]. After segmentation, T1ρ relaxation times were calculated for all voxels within each disc before the discs were rotated into a standard coordinate system and bisected in order to isolate the sub-region adjacent to each endplate. The central 40% of the disc in the anterior-posterior direction was used to represent the NP region [14]. The Pfirrmann grade of each disc was scored by a radiologist [T.L.] on T2-weighted images [17].

### Outcomes and statistical methods

The primary outcomes were the mean CEP T2* value in the central CEP, the mean BMFF in the vertebral body adjacent to each CEP, and the mean T1ρ value in the NP region adjacent to each CEP. Univariate linear regression was used to assess independent relationships between NP T1ρ (dependent variable), CEP T2*, and vertebral BMFF (independent variables). Logistic regression was used to assess the relationship between mean NP T1ρ and Pfirrmann grade. A mixed effects multivariable linear regression model with a random intercept accounting for multiple observations (from multiple spinal levels) per subject as well as age, sex, and BMI was used to assess the combined relationship between NP T1ρ, CEP T2*, and vertebral BMFF. Spinal level, and the interaction between level and CEP T2* were also included in the mixed effects model. Using the statistically significant predictors of NP T1ρ from the aforementioned multivariable model, a separate multivariable mixed effects linear regression model was created to compare the effects of CEP T2*, vertebral BMFF, and their interaction. Log transformations of the data were explored but not included since the untransformed data best fit the models based on assessment of residuals.

Finally, we used published regression data [6] relating CEP T2* with tissue compositional measures to estimate the distribution of GAG, water content, and collagen-to-GAG ratio in each CEP.

Statistical analyses were conducted in JMP Pro (16.0) and two-sided *p <* 0.05 was considered statistically significant.

## Results

Complete MRI data were successfully acquired from 80/84 prospectively enrolled patients. Following exclusion based on CEP damage (20 subjects) and orientation (110 endplates), 130 endplates from 60 patients were included in the final analysis (age = 40.0 ± 11.9 years, 28 (47%) female and 32 (53%) male, Table 1). There was a broad range of disc degeneration in the included levels: NP T1ρ ranged from 33.9–114.7 ms with mean ± SD = 62.4 ± 18.0 ms; Pfirrmann grades ranged from I–IV. NP T1ρ was significantly and negatively correlated with Pfirrmann grade (*p* < 0.0001), confirming that lower T1ρ values reflected more severe disc degeneration.

**Table 1.**
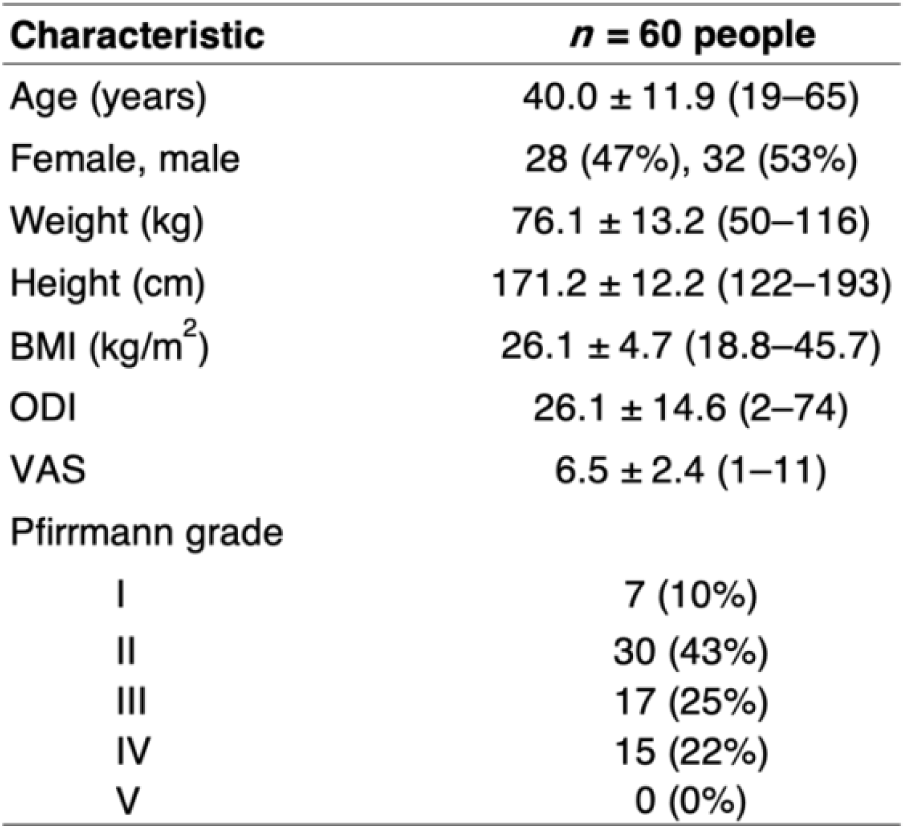
Patient demographic and clinical data from the *n =* 60 people included in the analysis. Data are presented as mean ± SD (min–max) or number (percent of total)

Results from the univariate regression analysis demonstrated that CEP T2* and vertebral BMFF were each independently significantly associated with NP T1ρ (*p* = 0.003 and 0.0001, respectively; Figure 2). These explanatory variables, as well as NP T1ρ, were significantly correlated with age (Table 2); after adjusting for age, sex, and BMI, NP T1ρ remained significantly associated with CEP T2* (*p* = 0.0001, Table 3) but not vertebral BMFF (*p =* 0.43). There was a statistically significant effect of level (*p <* 0.0001), indicating more severe disc degeneration at L5/S1 compared with L4/5 in these patients (mean NP T1ρ = 72.1 ms [95% CI: 66.7, 77.5] versus 60.5 ms [56.5, 64.5] for L4/5 versus L5/1, *p* < 0.0001). There was also a statistically significant interaction between level and CEP T2* (*p =* 0.0006), indicating that the relationship between CEP composition and disc degeneration was different at L4/5 compared with L5/S1 (NP T1ρ was more sensitive to variations in CEP T2* at L4/5 than at L5/S1).

**Fig. 2.**
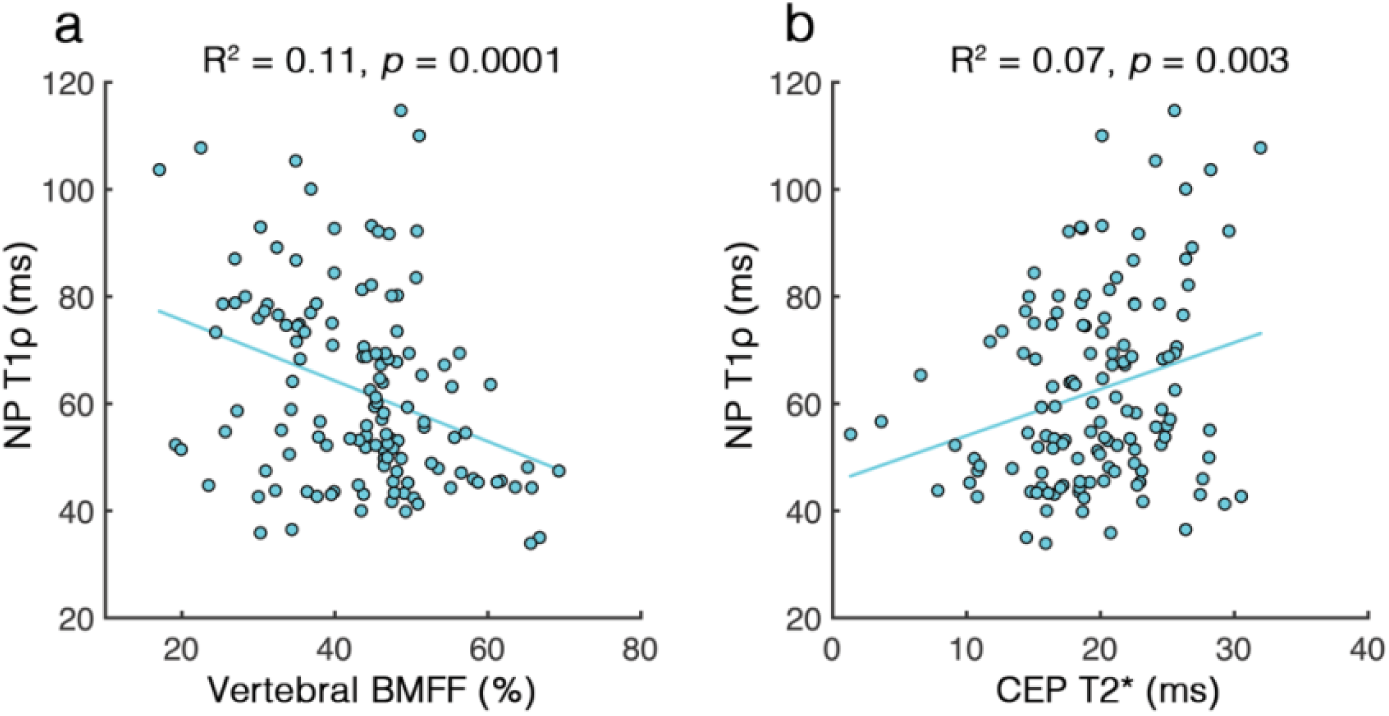
Scatter plots showing the independent relationships between NP T1ρ and (a) vertebral BMFF and (b) CEP T2*

**Table 2.** Pearson correlation coefficient quantifying the cross-correlations between NP T1ρ and the continuous explanatory variables

|  | **NP T1p** | **CEP T2\*** | **BMFF** | **Age** |
| --- | --- | --- | --- | --- |
| CEP T2* | 0.259 ** | - |  |  |
| BMFF | -0.333 *** | -0.160 | - |  |
| Age | -0.381 *** | -0.185 * | 0.599 *** | - |
| BMI | 0.043 | 0.339 *** | -0.049 | -0.100 |

\* $p < 0.05$
\*\* $p < 0.01$
\*\*\* $p \leq 0.0001$

**Table 3.** Parameter estimates (βi) and 95% confidence intervals generated from a mixed effects linear regression model accounting for multiple spinal levels per subject predicting NP T1ρ (outcome). Asterisk indicates two-sided *p* < 0.05

| Term | $\beta_i$ estimate | Lower 95% | Upper 95% | $p$-value |
| --- | --- | --- | --- | --- |
| CEP T2* | 1.06 | 0.53 | 1.59 | 0.0001* |
| Level | 5.80 | 3.76 | 7.84 | <.0001* |
| CEP T2* × Level | 0.83 | 0.37 | 1.30 | 0.0006 |
| Age | -0.45 | -0.83 | -0.06 | 0.0245* |
| Vertebral BMFF | -0.13 | -0.47 | 0.20 | 0.431 |
| Sex | 2.32 | -1.89 | 6.53 | 0.275 |
| BMI | -0.26 | -1.15 | 0.64 | 0.567 |

A second multivariable regression model included the significant predictors of NP T1ρ (age and level) plus CEP T2*, vertebral BMFF, and their interaction terms. Consistent with first multivariable model, results showed that CEP T2*, age, and spinal level were significant predictors of NP T1ρ while vertebral BMFF was not (Table 4). The interaction between CEP T2* and vertebral BMFF was not significant (*p* = 0.73), indicating the relationship between NP T1ρ and CEP T2* did not depend on vertebral BMFF.

**Table 4.**
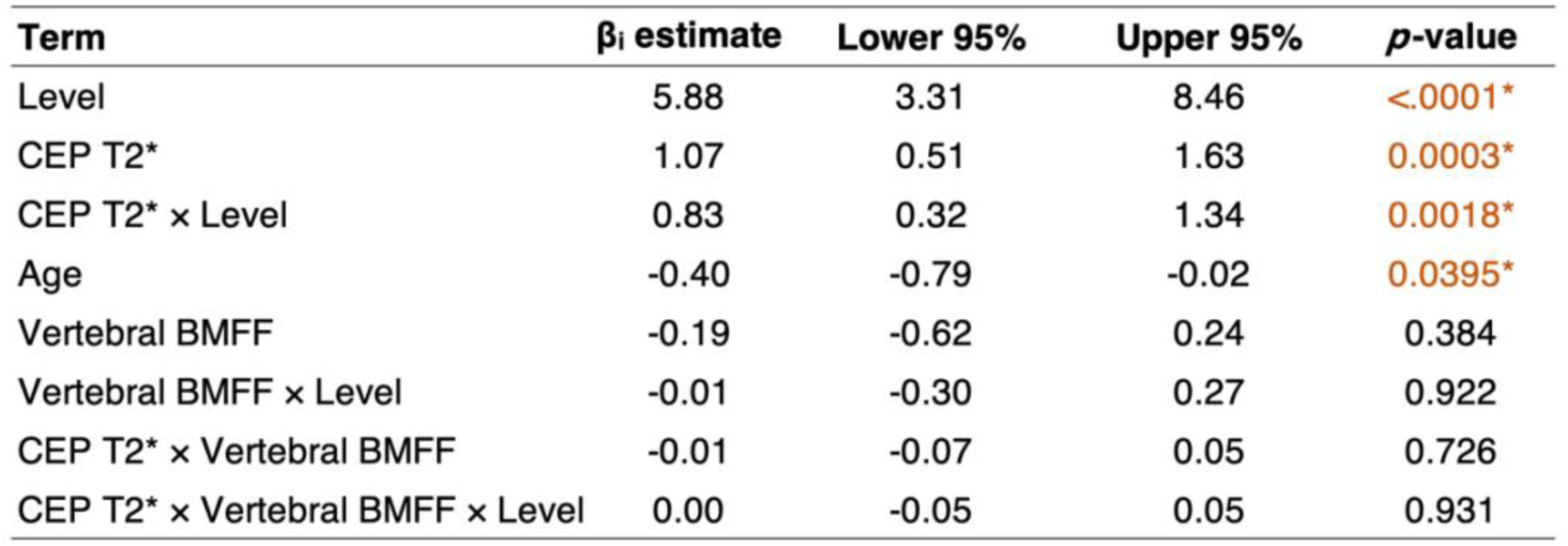
Parameter estimates (βi) and 95% confidence intervals generated from a mixed effects linear regression model accounting for multiple spinal levels per subject predicting NP T1ρ (outcome). Asterisk indicates two-sided *p* < 0.05

| Term | $\beta_i$ estimate | Lower 95% | Upper 95% | $p$-value |
| --- | --- | --- | --- | --- |
| Level | 5.88 | 3.31 | 8.46 | <.0001* |
| CEP T2* | 1.07 | 0.51 | 1.63 | 0.0003* |
| CEP T2* × Level | 0.83 | 0.32 | 1.34 | 0.0018* |
| Age | -0.40 | -0.79 | -0.02 | 0.0395* |
| Vertebral BMFF | -0.19 | -0.62 | 0.24 | 0.384 |
| Vertebral BMFF × Level | -0.01 | -0.30 | 0.27 | 0.922 |
| CEP T2* × Vertebral BMFF | -0.01 | -0.07 | 0.05 | 0.726 |
| CEP T2* × Vertebral BMFF × Level | 0.00 | -0.05 | 0.05 | 0.931 |

Based on the observed range of CEP T2* values along with published regression equations relating CEP T2* values to biochemical measurements [6], CEP GAG content was estimated to range from 58–95 µg/mg dry weight, water content from 49–66%, and collagen-to-GAG ratio from 4.8–10.0.

## Discussion

These results provide the first *in vivo* evidence that lumbar disc degeneration severity in patients with cLBP relates to compositional deficits in the CEP. We found that CEPs with lower T2* values—reflecting lower CEP hydration, lower GAG content, a higher ratio of collagen-to-GAG, and lower transport properties [4,6]—were associated with more severe disc degeneration both before and after accounting for patient age, sex, BMI, and multiple measurements per subject. Conversely, higher BMFF values in the adjacent vertebral body—reflecting less hematopoietic marrow and decreased perfusion [7–10,18]—were not associated with disc degeneration after accounting for covariates (namely, subject age). CEP composition strongly influences its permeability: CEPs with lower hydration and greater collagen-to-GAG ratios are less permeable than more hydrated and less dense CEPs [4]. Taken together, these collective findings suggest that CEP composition affects disc health by regulating nutrient and metabolite transport between the vertebral capillaries and the NP. We conclude that poor CEP composition plays a significant role in disc degeneration severity in patients with cLBP, and that CEP composition can affect disc health both with and without deficits in vertebral perfusion.

CEP composition and vertebral vascularity are known to be important factors in disc health, though their relative importance has remained unclear because prior studies have focused on each factor alone, neglecting their combined effects. For example, *in vitro* experiments have demonstrated that human CEP biochemical composition affects transport properties [19], with physiologic variations in the amounts of aggrecan, mineral, and collagen impacting nutrient diffusion to a degree sufficient to substantially hinder NP cell survival and function [4]. Deficits in nutrient supply have also been shown to reduce NP cell viability [5]. Those prior findings demonstrate the mechanisms by which CEP composition and vertebral vascularity may affect NP cells; our present findings corroborate the importance of CEP composition in the context of physiologic variations in vertebral vascularity. Unlike vertebral BMFF, CEP composition was a significant predictor of disc health after accounting for co-variates, which demonstrates the relative importance of CEP composition in disc degeneration in patients with cLBP.

Our finding that disc health strongly associates with CEP composition has implications for the clinical translation of emerging biologic therapies designed to regenerate the disc. The therapeutic potential of cell-based regenerative therapies likely depends on disc nutrient supply: mesenchymal stem cell chondrocytic differentiation, proliferation, and function depend on glucose and oxygen concentrations (the same nutrients vital to NP cell survival) and on matrix acidity (related to lactate production, an NP cellular metabolic waste product) [20]. Given the established effects of CEP composition on nutrient transport in combination with the data shown here, it follows that the efficacy of cell-based therapies may likewise depend on CEP composition. If so, CEP composition could be a possible diagnostic target to help identify patients with adequate CEP permeability to support increased cell density/activity in the NP, and/or a possible therapeutic target to enhance the disc’s regenerative potential, *e*.*g*. through enzymatic augmentation [21]. Non-invasive assessment of CEP composition with UTE MRI may thus provide a powerful tool to improve selection of patients and spinal levels likely to respond to anabolic treatment, helping to maximize the efficacy of novel biologic therapies.

The variations in biomarker values with age reported here are consistent with established age-associated biochemical changes in these tissues [18,22,23]. Lumbar CEP GAG and water contents in 51–67 year old subjects ranged from 43–185 µg/mg dry weight and 22–62%, respectively, in a previous study [24]. For patients with similar ages in the current study (49–65 year old), the estimated GAG and water content ranges (65–84 µg/mg dry weight and 52–63% water) are within the range previously published. A prior study using CSE-MRI [8] found that higher levels of BMFF were associated with more severe disc degeneration, consistent with the univariate regression results shown here. These findings collectively support the validity of our approach.

One strength of our study is the deployment of two recently developed neural networks [15,16] to perform automated tissue segmentation. This enabled rapid and unbiased evaluation of the MRI biomarkers, demonstrating the relevance of neural networks to a diverse class of problems and addressing the inefficiency and subjectivity associated with manual tissue segmentation. Notably, we also demonstrated the feasibility of non-invasively assessing CEP composition with a new MRI biomarker (mean CEP T2*) in a clinical population.

This study has several limitations. First, the cross-sectional study design precludes establishment of causality. However, prior research has demonstrated the causal effects of these same CEP compositional traits on NP cell survival and function [4], which provides a mechanistic explanation for the results shown here. A second limitation relates to the spatial resolution of the vertebral BMFF maps (voxel dimensions 1.0 × 1.0 × 4.0 mm), which does not capture subtle morphologic variations in endplate-specific vascularity which may affect nutrient supply [25]. However, the conversion from hematopoietic to fatty marrow has been shown to associate with vascular changes in both the femur and vertebral body and with age-associated changes in perfusion [9,10,18], suggesting that vertebral BMFF values likely provide an accurate measure of overall changes in vertebral vascularity.

In summary, we found that after adjusting for age, sex, and BMI, deficits in CEP composition—as indicated by lower T2* values—were associated with more severe disc degeneration. Conversely, lower vertebral vascularity—as indicated by higher vertebral BMFF values—was not associated with disc degeneration. We conclude that poor CEP composition plays a significant role in disc degeneration severity in patients with cLBP both with and without deficits in vertebral vascularity.

## Data Availability

Data produced in the present study may be available upon request at the discretion of the authors.

## Acknowledgements

The authors acknowledge Ronit Gupta for his help in data collection that contributed to the results shown here. Software tools facilitating data analysis were developed with support from NIH HEAL Initiative award UH2AR076724.

## Supplemental Material

### Detailed methods

MRI of the lumbar spine was performed on a GE 3T Discovery MR750 scanner using an 8-channel phased-array spine coil (GE Healthcare, Waukesha, WI). MRI data used for the purposes of this study were acquired using sagittal acquisitions from a multi-echo UTE Cones sequence, a chemical shift encoding-based water-fat sequence, a T1ρ mapping sequence, and standard clinical fast spin-echo sequences with T1- and T2-weighting. The sequence details are as follows:

1. 1. Multi-echo 3D UTE Cones sequence: echo time (TE) = 0.24, 5.2, 10.2, 15.2, 20.2, 25.2 ms; repetition time (TR) = 36 ms; field-of-view (FOV) = 28 cm; flip angle = 15°; in-plane resolution = 0.5 mm; slice-thickness = 3 mm (interpolated to 1 mm).
2. 2. Six-echo water-fat sequence consisting of a 3D spoiled gradient-recalled echo (SPGR) sequence with iterative decomposition of water and fat with echo asymmetry and least-squares estimation (IDEAL) reconstruction: TE = 2, 3, 4, 5, 6, 7 ms; TR = 6.2 ms; FOV = 28 cm; flip angle = 3°; in-plane resolution = 1.0 mm; slice thickness = 4 mm, receiver bandwidth = 83.3 kHz.
3. 3. 3D T1ρ mapping sequence consisting of a magnetization-prepared angle-modulated partitioned k-space SPGR sequence: spin-lock time (TSL) = 0, 10, 40, and 80 ms; TR = 5.2 ms; FOV = 20 cm; flip angle = 60°; in-plane resolution = 0.78 mm ; slice thickness = 8 mm; spin-lock frequency = 300 Hz.
4. 4. Standard clinical fast spin-echo with T1- and T2-weighting: TE = 15, 60 ms (T1-, T2-weighted, respectively); TR = 511, 4877 ms; echo train length (ETL) = 24, 4; FOV = 26 cm; in-plane resolution = 0.5 mm; slice thickness = 4 mm; receiver bandwidth = 50 kHz.

## Works cited

[1] Urban J, Fairbank J. Current perspectives on the role of biomechanical loading and genetics in development of disc degeneration and low back pain; a narrative review. J Biomech. 2020;102:109573. https://doi.org/10.1016/j.jbiomech.2019.109573

[2] Binch A, Fitzgerald J, Growney E, et al. Cell-based strategies for IVD repair: clinical progress and translational obstacles. Nat Rev Rheumatol. 2021;17:158–75. https://doi.org/10.1038/s41584-020-00568-w

[3] Ju D, Kanim L, Bae H. Is there clinical improvement associated with intradiscal therapies? A comparison across randomized controlled studies. Glob Spine J. 2020;1–9. https://doi.org/10.1177/2192568220963058

[4] Wong J, Sampson S, Bell-Briones H, et al. Nutrient supply and nucleus pulposus cell function: effects of the transport properties of the cartilage endplate and potential implications for intradiscal biologic therapy. Osteoarthr Cartil. 2019;27(6):956–64. https://doi.org/10.1016/j.joca.2019.01.013

[5] Horner H, Urban J. 2001 Volvo award winner in basic science studies: Effect of nutrient supply on the viability of cells from the nucleus pulposus of the intervertebral disc. Spine (Phila Pa 1976). 2001;26(23):2543–9. https://doi.org/10.1097/00007632-200112010-00006

[6] Fields A, Han M, Krug R, et al. Cartilaginous end plates: quantitative MR imaging with very short echo times-orientation dependence and correlation with biochemical composition. Radiology. 2015;274(2):482–9. https://doi.org/10.1148/radiol.14141082

[7] Gee C, Nguyen J, Marquez C, et al. Validation of bone marrow fat quantification in the presence of trabecular bone using MRI. J Magn Reson Imaging. 2015;42(2):539–44. https://doi.org/10.1002/jmri.24795

[8] Krug R, Joseph G, Han M, et al. Associations between vertebral body fat fraction and intervertebral disc biochemical composition as assessed by quantitative MRI. J Magn Reson Imaging. 2019;50(4):1219–26. https://doi.org/10.1002/jmri.26675

[9] Chen W, Shih T, Chen R, et al. Vertebral bone marrow perfusion evaluated with dynamic contrast-enhanced MR imaging: significance of aging and sex. Radiology. 2001;220(1):213–8. https://doi.org/10.1148/radiology.220.1.r01jl32213

[10] Bluemke D, Petri M, Zerhouni E. Femoral head perfusion and composition: MR imaging and spectroscopic evaluation of patients with systemic lupus erythematosus and at risk for avascular necrosis. Radiology. 1995;197(2):433–8. https://doi.org/10.1148/radiology.197.2.7480688

[11] Auerbach J, Johannessen W, Borthakur A, et al. In vivo quantification of human lumbar disc degeneration using T1ρ-weighted magnetic resonance imaging. Eur Spine J. 2006;15(Suppl. 3):338–44. https://dx.doi.org/10.1007%2Fs00586-006-0083-2

[12] Deyo R, Dworkin S, Amtmann D, et al. Report of the NIH task force on research standards for chronic low back pain. J Pain. 2014;15(6):569–85. https://doi.org/10.1016/j.jpain.2014.03.005

[13] Gurney P, Hargreaves B, Nishimura D. Design and analysis of a practical 3D cones trajectory. Magn Reson Med. 2006;55(3):575–82. https://doi.org/10.1002/mrm.20796

[14] Wang L, Han M, Wong J, et al. Evaluation of human cartilage endplate composition using MRI: spatial variation, association with adjacent disc degeneration, and in vivo repeatability. J Orthop Res. 2020;39(7):1470–8. https://doi.org/10.1002/jor.24787

[15] Zhou J, Damasceno P, Chachad R, et al. Automatic vertebral body segmentation based on deep learning of Dixon images for bone marrow fat fraction quantification. Front Endocrinol (Lausanne). 2020;11(September):612. https://doi.org/10.3389/fendo.2020.00612

[16] Iriondo C, Pedoia V, Majumdar S. Lumbar intervertebral disc characterization through quantitative MRI analysis: an automatic voxel-based relaxometry approach. Magn Reson Med. 2020;84(3):1376–90. https://doi.org/10.1002/mrm.28210

[17] Pfirrmann C, Metzdorf A, Zanetti M, et al. Magnetic resonance classification of lumbar intervertebral disc degeneration. Spine (Phila Pa 1976). 2001;26(17):1873–8. https://doi.org/10.1097/00007632-200109010-00011

[18] De Bisschop E, Luypaert R, Louis O, et al. Fat fraction of lumbar bone marrow using in vivo proton nuclear magnetic resonance spectroscopy. Bone. 1993;14(2):133–6. https://doi.org/10.1016/8756-3282(93)90239-7

[19] Roberts S, Urban J, Evans H, et al. Transport properties of the human cartilage endplate in relation to its composition and calcification. Spine (Phila Pa 1976). 1996;21(4):415–420. https://doi.org/10.1097/00007632-199602150-00003

[20] Huang Y, Leung V, Lu W, et al. The effects of microenvironment in mesenchymal stem cell-based regeneration of intervertebral disc. Spine J. 2013;13(3):352–62. https://doi.org/10.1016/j.spinee.2012.12.005

[21] Dolor A, Sampson S, Lazar A, et al. Matrix modification for enhancing the transport properties of the human cartilage endplate to improve disc nutrition. PLoS One. 2019;14(4):e0215218. https://doi.org/10.1371/journal.pone.0215218

[22] Johannessen W, Auerbach J, Wheaton A, et al. Assessment of human disc degeneration and proteoglycan content using T1ρ-weighted magnetic resonance imaging. Spine (Phila Pa 1976). 2006;31(11):1253–1257. https://dx.doi.org/10.1097%2F01.brs.0000217708.54880.51

[23] Antoniou J, Goudsouzian N, Heathfield T, et al. The human lumbar endplate: evidence of chnages in biosynthesis and denaturation of the extracellular matrix with growth, maturation, aging, and degeneration. Spine (Phila Pa 1976). 1996;21(10):1153–1161. https://doi.org/10.1172/jci118884

[24] Fields A, Rodriguez D, Gary K, et al. Influence of biochemical composition on endplate cartilage tensile properties in the human lumbar spine. J Orthop Res. 2014;32(2):245–52. https://dx.doi.org/10.1002%2Fjor.22516

[25] Gullbrand S, Peterson J, Mastropolo R, et al. Drug-induced changes to the vertebral endplate vasculature affect transport into the intervertebral disc in vivo. J Orthop Res. 2014;32(12):1694–700. https://doi.org/10.1002/jor.22716

